# Proliferative History Is a Novel Driver of Clinical Outcome in Splenic Marginal Zone Lymphoma

**DOI:** 10.1101/2024.01.16.24301320

**Authors:** Helen Parker, Amatta Mirandari, Carolina Jaramillo Oquendo, Martí Duran-Ferrer, Benjamin Stevens, Lara Buermann, Harindra E. Amarasinghe, Jaya Thomas, Latha Kadalayil, Louise Carr, Shama Syeda, Methusha Sakthipakan, Marina Parry, Zadie Davis, Neil McIver-Brown, Aliki Xochelli, Sarah Ennis, Lydia Scarfo, Paolo Ghia, Christina Kalpadakis, Gerassimos Pangalis, Davide Rossi, Simon Wagner, Matthew Ahearne, Marc Seifert, Christoph Plass, Dieter Weichenhan, Eva Kimby, Lesley-Ann Sutton, Richard Rosenquist, Francesco Forconi, Kostas Stamatopoulos, Marta Salido, Ana Ferrer, Catherine Thieblemont, Viktor Ljungström, Rose-Marie Amini, David Oscier, Renata Walewska, Matthew J.J. Rose-Zerilli, Jane Gibson, José Ignacio Martín-Subero, Christopher Oakes, Dean Bryant, Jonathan C Strefford

## Abstract

The epiCMIT (epigenetically-determined Cumulative MIToses) mitotic clock traces B-cell mitotic history via DNA methylation changes in heterochromatin and H3K27me3-containing chromatin. While high scores correlated with poor outcomes in CLL and MCL, its prognostic significance in SMZL remains unknown. Derived from 142 SMZL cases using DNA methylation microarrays, epiCMIT values were correlated with genomic, transcriptomic, and clinical data. EpiCMIT as a continuous variable was significantly higher in females (*p*=0.02), patients with IGHV1-2*04 allele usage (*p*<0001), intermediate IGHV somatic hypermutation load (97-99.9% identity, *p*=0.04), elevated mutational burden (25 vs. 17 mut/Mb, *p*=0.001), driver gene mutations [*KLF2* (*p*<0.001), *NOTCH2* (*p*<0.01), *TP53* (*p*=0.01), *KMT2D* (*p*<0.001)], and del(7q) (*p*=0.01). Negative correlation between epiCMIT and telomere length (r=-0.29 *p*<0.001) supported the association between cumulated proliferation and telomere attrition. While univariate analysis highlighted epiCMIT as robust predictor of shorter treatment-free survival (TFS), multivariate analysis confirmed epiCMIT as an independent marker for shorter TFS. In summary, our matched multi-omic datasets facilitate the clinico-biological characterization of SMZL and introduces epiCMIT as a strong prognostic marker, identifying high-risk patients and predicting reduced treatment-free survival, hence providing a new tool for risk-adapted patient management.

## Introduction

Splenic marginal zone lymphoma (SMZL) is a rare B-cell malignancy in adults. Whilst having morphological and biological similarities to other marginal zone lymphomas, current classifications of lymphoid tumors group SMZL with other primary splenic B cell lymphomas, which involve the spleen, bone marrow and frequently the blood and perihilar but not peripheral lymph nodes (1–4). SMZL has a characteristic splenic histology and is associated with recurring, albeit not pathognomonic, genomic abnormalities which include 7q deletions (∼40% cases) (5), *KLF2* (20–30%), *NOTCH2* (10–25%), *TP53* mutations (10–15%) (6–10) and a biased IGHV gene repertoire with usage of IGHV1-2*04 in 30% of cases (6, 11, 12). Diagnosis is usually based on a combination of clinical features, lymphocyte morphology, bone marrow histology and immunophenotype in most cases who do not undergo splenectomy. Despite a generally favorable prognosis and a median survival of 10-15 years, clinical outcome is heterogeneous. 10% of patients will present with a more aggressive disease, a further 20% will go on to develop progressive disease requiring treatment, and histological transformation to large-cell lymphoma occurs in 10-20% of these cases (13, 14).

With the emergence of new targeted therapies and the potential for immune therapies, there is a need for biomarkers to guide both the nature and timing of treatment. To this end, Bonfiglio *et al* (15) have recently identified four genetic clusters and two distinct splenic microenvironment profiles with prognostic significance among a large cohort of SMZL patients who had undergone splenectomy as first line therapy. Those cases with an inflamed splenic microenvironment and a genetic cluster enriched for mutations in *KLF2*, NF-kB and/or NOTCH pathways, had a relative survival at 10 years of 70.8% compared with matched controls. Turning from genetics to epigenetics, a study of genome-wide DNA-promoter methylation, Arribas and colleagues identified two clusters of SMZL patients with different degrees of promoter methylation (16) and clinical outcome. The high methylation cluster was associated with a poorer overall survival and with IGHV1-2*04 gene usage, *NOTCH2* mutations, 7q deletions and histologic transformation. However, the broader impact of DNA methylation, transcending its traditional gene regulatory role, warrants closer examination; hypo-methylation in low CpG-content heterochromatin and hyper-methylation in high CpG-content polycomb regions accumulate during cell division. These changes reflect the mitotic history of B cell tumors, not only following malignant transformation but also during prior normal B-cell maturation and can be assessed by the recently developed epigenetically-determined Cumulative MIToses (epiCMIT) score. Whilst epiCMIT correlates with outcomes, recurrent gene mutations, genome complexity, mutational signatures, and higher expression of cell proliferation-related genes in chronic lymphocytic leukemia (CLL) and mantle cell lymphoma (MCL) (17), no such studies have been performed in SMZL.

This study employed genome-wide DNA methylation data on a large, international cohort of SMZL, with matched mutational, copy-number, telomere length and clinical data, to address the clinical and biological importance of the epiCMIT score in SMZL. In doing so, we show that the degree of proliferative history significantly associated with key genomic, immunogenetic and biological disease features. Importantly, epiCMIT was a marker of treatment free survival (TFS) in both univariate and multivariate survival models, suggesting clinical utility in the risk stratification of SMZL patients.

## Methods

### Patient Cohort

Our cohort comprises 142 SMZL patients, all meeting established diagnostic criteria. Informed consent was obtained from all patients in accordance with the Helsinki declaration, and the study was approved by our regional and local ethics committees of each participating institution. Sampling was performed as close to diagnosis as possible, with a median time between diagnosis and sampling of 4 months (range 0-49 months). Patients were sampled pre-treatment, or at the requirement for treatment in splenectomized cases. Prior to DNA extraction (DNeasy blood and tissue kit, Qiagen, Hilden, Germany), the CD19+/CD45+ SMZL cells were purified from the peripheral blood (n=100) or spleen (n=42) using the EasySep Human B Cell enrichment kit without CD43 depletion (StemCell Technologies, Cambridge, UK). Tumor purity of greater than 85% was confirmed in all cases by flow cytometry (CD19).

### DNA Methylation analysis

142 DNA samples were processed, as previously reported (18, 19) using the Illumina Infinium Human Methylation 450 BeadChip (n=111) and Illumina Infinium Methylation EPIC BeadChip (n=31) (Illumina, Hayward, CA, USA), according to manufacturer’s instructions, at the Genomics and Proteomics Core Facility of the DKFZ (Heidelberg, Germany). Data processing was performed using RnBeads v2.93 (RRID:SCR_010958) (**Supplementary methods**). Conumee (20) was used to produce profiles of copy-number alterations (CNA); genomic complexity (GC) was defined as the presence of ³3 CNA. EpiCMIT-hypo and -hyper scores were calculated as per Duran-Ferrer *et al* (17) using the Estimate.epiCMIT function published by the authors. For each patient, the highest score from the epiCMIT-hypo and -hyper scores was selected to derive a unique epiCMIT value (termed epiCMIT). Cases were assigned to the Arribas-High (AH, n=28) or -Low (AL, n=114) methylation subgroup by clustering beta values using Euclidean distance and complete linkage. Tumor purity estimates, calculated from DNA methylation data as previously published (21), confirmed FACS based purity scores in all samples (**Fig. S1)**.

### Mutation detection with targeted re-sequencing

133 DNA samples were analyzed with a bespoke Haloplex Target Enrichment panel (Agilent Technologies, Santa Clara, CA, USA), designed to enrich for 62 genes and additional genomic regions, selected for their clinical relevance to SMZL and other B-cell malignancies (**Table S1**), as previously described (10). High confidence variants were identified using a customized bioinformatics pipeline and filtering strategy (**Supplementary methods**). As described by Bonfiglio *et al* (15), the presence of somatic mutations in a set of 14 genes can be used to assign SMZL patients into two prominent clusters termed NNK (NF-Kb, NOTCH and KLF2 modules) and DMT (DNA damage response, MAPK and TLR modules). Except for *PTPN11*, used to assign DMT, all genes used by the authors in this classification were included on our panel and were utilized to classify our patients as NNK (n=52), DMT (n=35) or unclassified/other (n=55).

### Telomere length analysis

Telomere length (TL) relative to a standard reference sample (K562 cell line, ATCC® CCL-243TM) was determined in 114 patients using monochrome multiplex PCR (MMQ-PCR) as previously described (22). Absolute TL (kb) was extrapolated, using linear regression, from 82 CLL cases with STELA data (**Supplementary methods**). Patients were classified as having long or short telomeres based on the median length (3.12 kb).

### Whole – genome and transcriptome sequencing

A sub-cohort of twenty-three patients, with diverse epiCMIT scores (range:0.37-0.90) and viable cells available, were processed with WGS (n=23/23) and mRNA-Seq (n=15/23). For WGS, DNA was extracted from tumor B-cells (Qiagen DNeasy Blood and Tissue Kit) and matched saliva (Oragene DNA Saliva kit, DNA Genotek, Ottawa, Canada), prior to library preparation and sequencing at the SNP&SEQ Technology Platform, Science for Life Laboratory at Uppsala University, Sweden. Data analysis was performed according to the GATK Best Practices Workflow (23), with bespoke parameters (**Supplementary methods**). Visualization of SNV calling data from .maf files was performed in R v4.3.1 (RRID:SCR_001905), using MAFTools v2.10.0 (RRID:SCR_024519) (23) and ggplot2 v3.3.5 packages (RRID:SCR_014601). The coding tumor mutational burden (TMB) was assessed using MAFTools v2.10.0 and was defined as the burden of mutations per Mb, within the coding regions of the genome. mRNA sequencing was performed using the NEBNext Poly(A) mRNA Magnetic Isolation Module (New England Biolabs, Massachusetts, USA) and sequenced on the Illumina NovaSeq6000. mRNA-seq analysis was performed as previously reported (18). Briefly, mRNA fasta files were aligned to the hg38 Reference genome using STAR aligner v2.7.10b (RRID:SCR_004463) and read counts were calculated through HTseq-count (RRID:SCR_011867) against Gencode GRCh38.p14 v44. Differential gene expression analysis was conducted using EdgeR v3.42.4 (RRID:SCR_012802) against epiCMIT as a continuous variable using likelihood ratio tests (significance; FDR p<0.05 with Benjamini-Hochburg correction). GSEA was performed against the Hallmark, KEGG and immune gene sets obtained from MSigDBv2023.1Hs (24–27). All analytical processes were conducted using R v4.3.1 (RRID:SCR_001905).

### Statistics

Relationships between variables, that were observed in at least 5% of cases (n=7), were compared using the Fisher’s exact and Mann-Whitney U tests (significance level *p*<0.05). Differences in survival measures between subgroups were tested with the log-rank test. Multivariate Cox Proportional Hazard models were generated for TFS using stepwise backwards elimination (**Supplementary methods**). All analyses were performed in R v3.6.1 (RRID:SCR_001905).

## Results

### Cohort overview

The overview of our cohort and methodological approaches is depicted in **Fig 1A**. Baseline clinico-biological characteristics of the 142 SMZL cases are shown in **Table 1**. 66 and 76 patients were male and female, respectively, with a median age of diagnosis of 69 years (range: 35-88 years). The cohort has a median follow-up of 5.75 years (range; 0-24), and includes 102 patients that have received treatment, 18 that have transformed, and 31 that have died. Splenectomy was the predominant first-line therapy (33.8% cases), followed by rituximab, either as a monotherapy or in combination with chemotherapy (29.5%). Subsequent treatment was necessary in 14.7% of patients with rituximab being the preferred choice in 71.4% cases.

**Figure 1.**
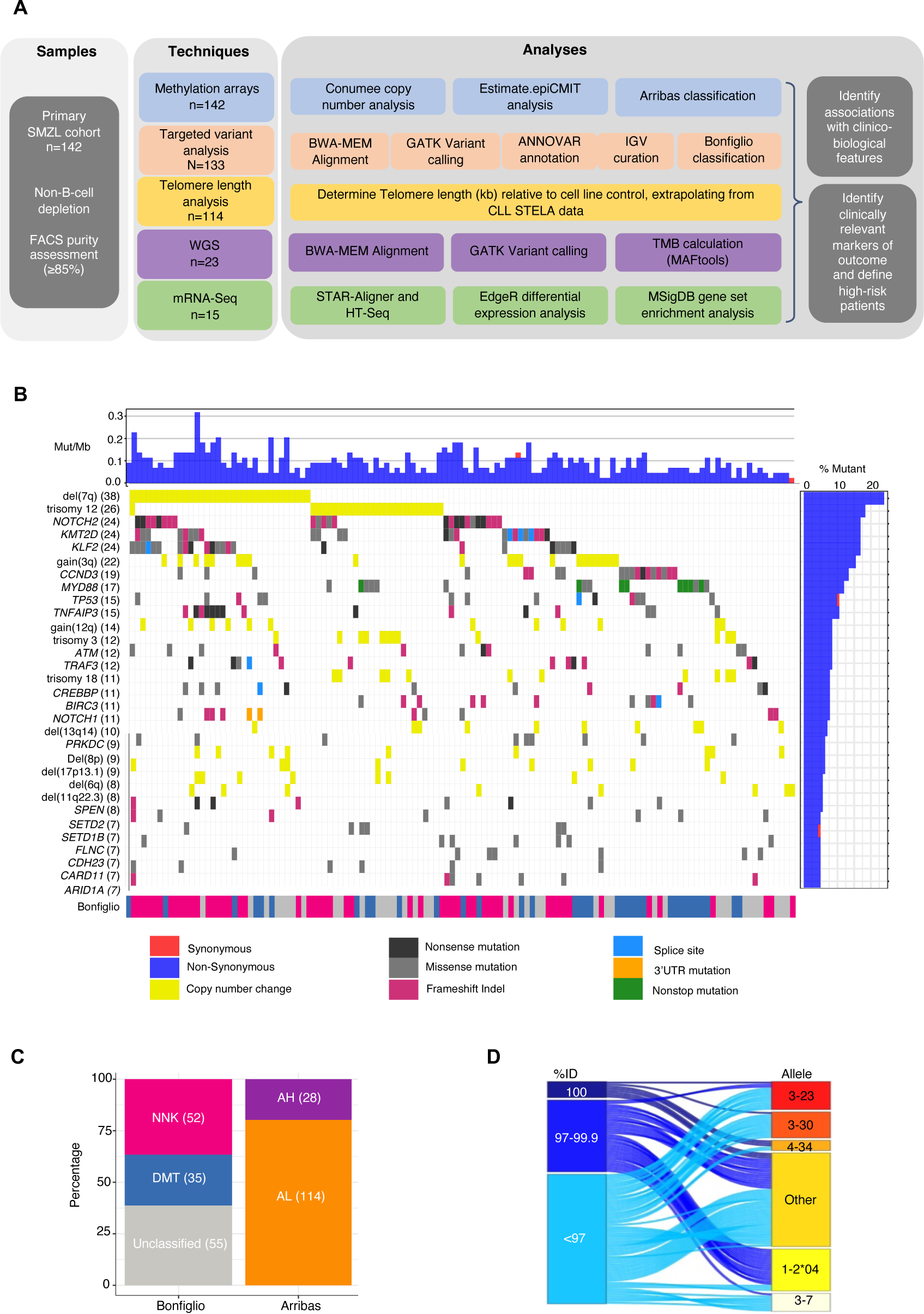
Project outline and overview of cohort composition. **A** Consort diagram showing the experimental workflow and key analytical processes applied to the data. **B** Waterfall plot showing the recurrently mutated genes and copy-number changes in SMZL, hierarchically ordered by aberration frequency (vertical bar chart, right and number ()). Mutational burden and mutation types are identified by the top panel and key, respectively. The bottom panel shows the Bonfiglio classification for each patient in pink (NNK), blue (DMT) and grey (unclassified). **C** Stacked bar chart showing the percentage of cases assigned the Bonfiglio classification of NNK, DMT or unclassified (15), and the Arribas classification of AH or AL. Number of cases are shown () (16). **D** Sankey plot showing the relationship between the IGHV somatic hyper-mutational load and the heavy chain allele usage (n= 107, unknown status=35).

**Table 1.** Key clinico-biological characteristics of the SMZL cohort.

| Variable (no. patients with variable data) | % of patients (numbers) |
| --- | --- |
| Female | 54% (76/142) |
| Age at diagnosis - median in years [range] | 67 [35-88] |
| Patients treated including splenectomy | 72% (102/142) |
| Patients transformed to large cell lymphoma | 13% (18/69) |
| Patients died | 22% (31/137) |
| OS - median in months [range] (128/142) | 63.0 [0-293] |
| TTFT- median in months [range] (136/142) | 10.0 [0-169] |
| HPLL/ ABC score (89/142) |  |
| A | 65.2% (58/89) |
| B | 34.8% (31/89) |
| IGHV1-2*04 usage (120/142) | 19% (27/120) |
| IGHV SHM load (110/142) |  |
| 100% | 5.6% (8/110) |
| 97-99.9% | 25.4% (36/110) |
| <97% | 46.5% (66/110) |
| *KLF2* mutations | 17% (24/133) |
| *KMT2D* mutations | 17% (24/133) |
| *NOTCH2* mutations | 17% (24/133) |
| *TP53* mutations | 11% (15/133) |
| Bonfiglio NNK classification | 36.6% (52/133) |
| Presence of Del 7q | 24% (34/142) |
| Complex genome ($\geq 3$ CNA) | 22.5% (32/142) |
| Telomere length (Kb) - median [range] | 3.12 [2.38-7.57] |
| AH classification | 20% (28/142) |
| EpiCMIT- median [range] | 0.74 [0.37-0.91] |

Our resequencing results were consistent with previously published data (6–10, 28–30); *NOTCH2*, *KMT2D* and *KLF2* were the most frequently mutated genes, each present in 24 patients (17%). In total, 341 high-confidence variants (mean 2.4, range 0-10 per patient) were identified in 44 genes with 20/44 genes recurrently mutated in >5% of the cohort (range 5-17%) (**Fig 1B, Table S2**). Seventeen *MYD88* mutations were identified with 47% mapping to the p.L265 hotspot. At least one CNA (range=0-14) was observed in 104/142 patients with 32 patients (22.5%) harboring a complex genome. The most prevalent CNAs were del(7q) (23.9%), trisomy 12 (18.3%), gain(3q) (15.5%) and gain(12q) (8.5%) (**Fig 1B and Fig S2A**). While chromosome 7q deletion breakpoints were heterogenous, a 1.75 Mb minimally deleted region (MDR) (128-57-130.32 Mb) was identified, that encompassed 24 coding genes, including *FLNC,* and 12 non-coding RNAs (**Fig S2B**). Five additional minimally deleted or gained regions were identified on chromosomes 3q, 8p, 12q, 13q and 17p (**Table S3**). Patients in our cohort were assigned a mutation-based classification; NNK (n=52) or DMT (n=35) using 7/7 and 6/7 key genes identified by Bonfiglio *et al* (15), respectively. Patients missing mutation data, or that had no mutations in the 13 genes, were designated ‘unclassified’ (n=55). Consistent with the authors findings, NNK patients were associated with IGHV1-2*04 (*p*=0.008) and del(7q) (*p*=0.01) (**Fig 1B**), whilst DMT patients were depleted for both. Furthermore, patient categorization into AH (Arribas-High) (n=28) and AL (Arribas-Low) (n=114) promotor methylation groups was executed, using 86 of the top 100 most variable CpGs identified by Arribas and colleagues (**Fig S3, Table S4**) (16). In line with their findings, patients classified as AH were characterized by the presence of IGHV1-2*04 (*p*=0.003), del(7q) (*p*=0.04), and *NOTCH2* mutations (*p*=0.02). All patients classified as AH were also classified as NNK (**Fig1C**). The most frequent IGHV genes were IGHV1-2 (27/142, 19%), IGHV3-23 (16/142, 11%), IGHV3-30 (14/142, 9%), IGHV3-7 (9/142, 6%) and IGHV4-34 6/142, 4%), with all IGHV1-2 cases harboring the *04 allele. The IGHV gene somatic hypermutation (SHM) status ranged from 88.9-100% identity to germline. The IGHV1-2*04 allele was associated with intermediate SHM load (97-99.9% germline identity, *p*=<0.001, 70% of cases) and female sex (*p*=<0.001, 81.5% cases). The presence of 100% germline identity (8/142, 5.6%) was mutually exclusive with IGHV1-2*04 but was associated with IGHV4-34 usage (*p*=0.002). IGHV3-23, IGHV3-30 and IGHV3-7 cases were enriched for high SHM load (<97% identity, *p*=<0.001) (**Fig 1D)**.

### Levels of proliferative history are highly variable in SMZL and associate with clinico-biological features

First, for each of our SMZL cases (n=142), we calculated the epiCMIT proliferative history score, a metric determined from methylation levels of CpGs that were observed in a previous study of mature B-cell tumors to become hypermethylated (epiCMIT-hyper) or hypomethylated (epiCMIT-hypo) with cumulative proliferation (17). The highest score obtained for epiCMIT-hyper or epiCMIT-hypo per sample was used as the epiCMIT score per patient (mean: 0.72, range: 0.37-0.91), reflecting the history of mitotic cell divisions pre- and post-transformation (**Fig 2A)**. We observed variable epiCMIT-hyper (mean: 0.6, range: 0.29-0.91) and epiCMIT-hypo (mean: 0.71, range: 0.37-0.90) values in our cohort (**Fig 2Bi**), and a strong positive correlation between them in SMZL (**Fig 2Bii**), as has been previously observed in MCL, CLL, DLBCL (17). The distribution of EpiCMIT values derived from splenic material or peripheral blood (from unpaired samples) did not significantly differ (p=0.15).

**Figure 2.**
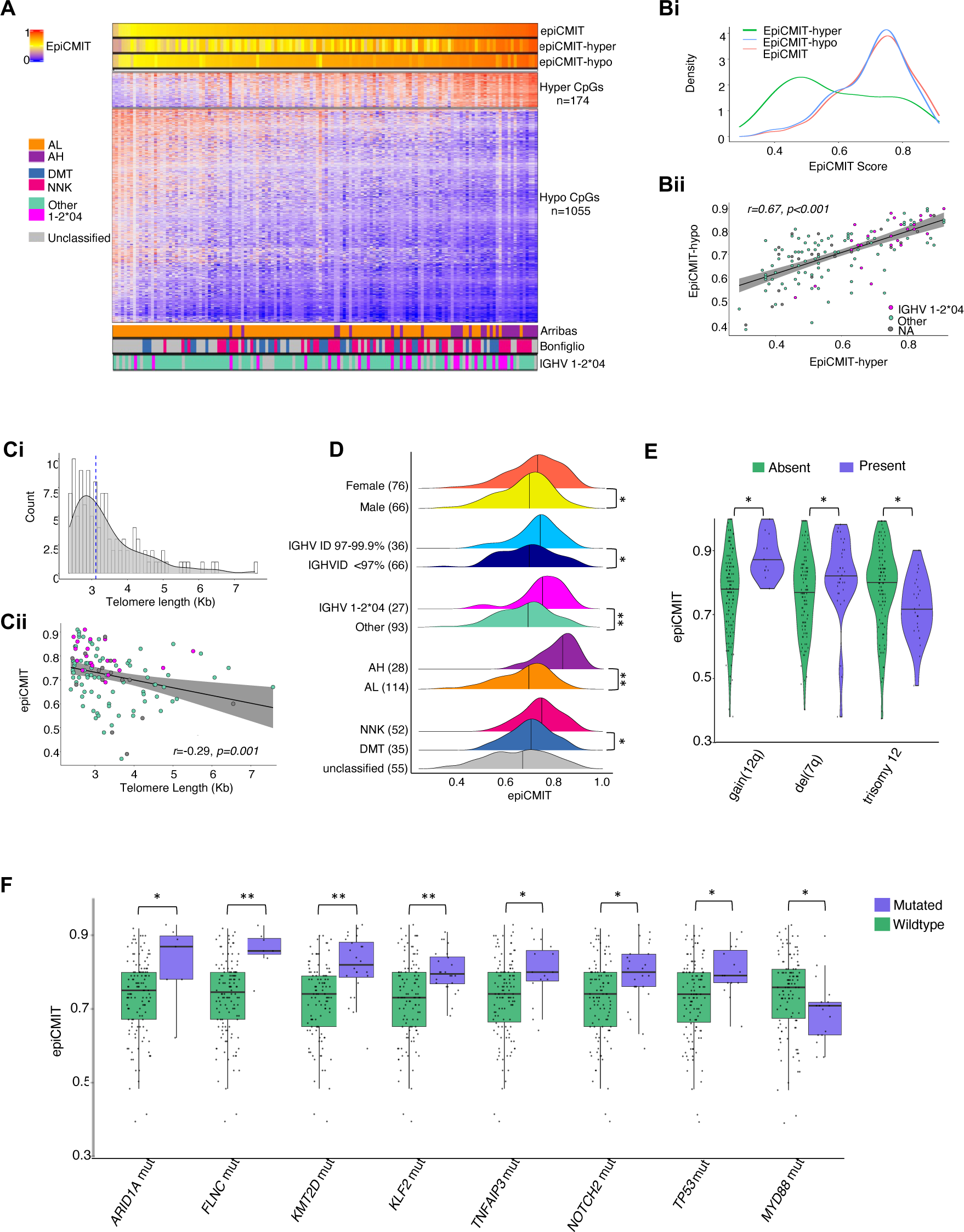
EpiCMIT and the association with clinico-biological features. **A** Heatmap showing the methylation status of 174 and 1055 CpGs used to calculate the epiCMIT-hyper and -hypo scores, respectively, with each patient represented on the *x* axis. The top panel of horizontal bars uses a graduated color scale to represent the increasing epiCMIT score. The bottom panel show the Arribas and Bonfiglio based classifications for each patient (15, 16), and the IGHV allele usage. **Bi** Density plot showing the distribution of epiCMIT-hyper, -hypo, and epiCMIT scores (green, blue and pink, respectively). **Bii** Scatterplot showing the positive correlation (p=<0.001) between increasing epiCMIT -hyper and -hypo scores. Points are colored purple or pink to indicate IGHV 1-2*04 or other usage, respectively (grey=data NA). The linear regression model is depicted by the black line, and the grey area indicates the confidence intervals **Ci.** Histogram showing the frequency and distribution of telomere length(kb) in our cohort, with the median length of 3.1kb identified by the blue dotted line. **Cii** Scatterplot showing the negative correlation between (p=0.001) high epiCMIT score and telomere length. **D** Ridgeplots showing the distribution of epiCMIT scores for variables exhibiting statistically significant differences between two status groups. Higher scores were observed in patients assigned NNK and AH status, females, and those with IGHV 1-2*04 usage and intermediate SHM loads (IGHV ID 100% not plotted (n=8)). The number of cases with each variable is shown (). **E** Violin plots showing the CNA with significant differences in epiCMIT scores between two status groups. Higher scores were observed in patients with gain(12q) and del(7q), whereas patients with trisomy 12 had lower scores. **F** Box and whisker plot showing the gene mutations associated with significantly different epiCMIT score between two status groups. Whilst patients with *ARID1A, FLNC, KMT2D, KLF2, TNFAIP3, NOTCH2* and *TP53* mutations were associated with a higher score, *MYD88* was associated with a lower score. Mann-Whitney U test with Benjamini-Hochberg correction was used to determine statistical significance (p=0.05). * and ** indicate p-values of *p*=<0.05 and *p*=<0.001, respectively.

**Figure 3.**
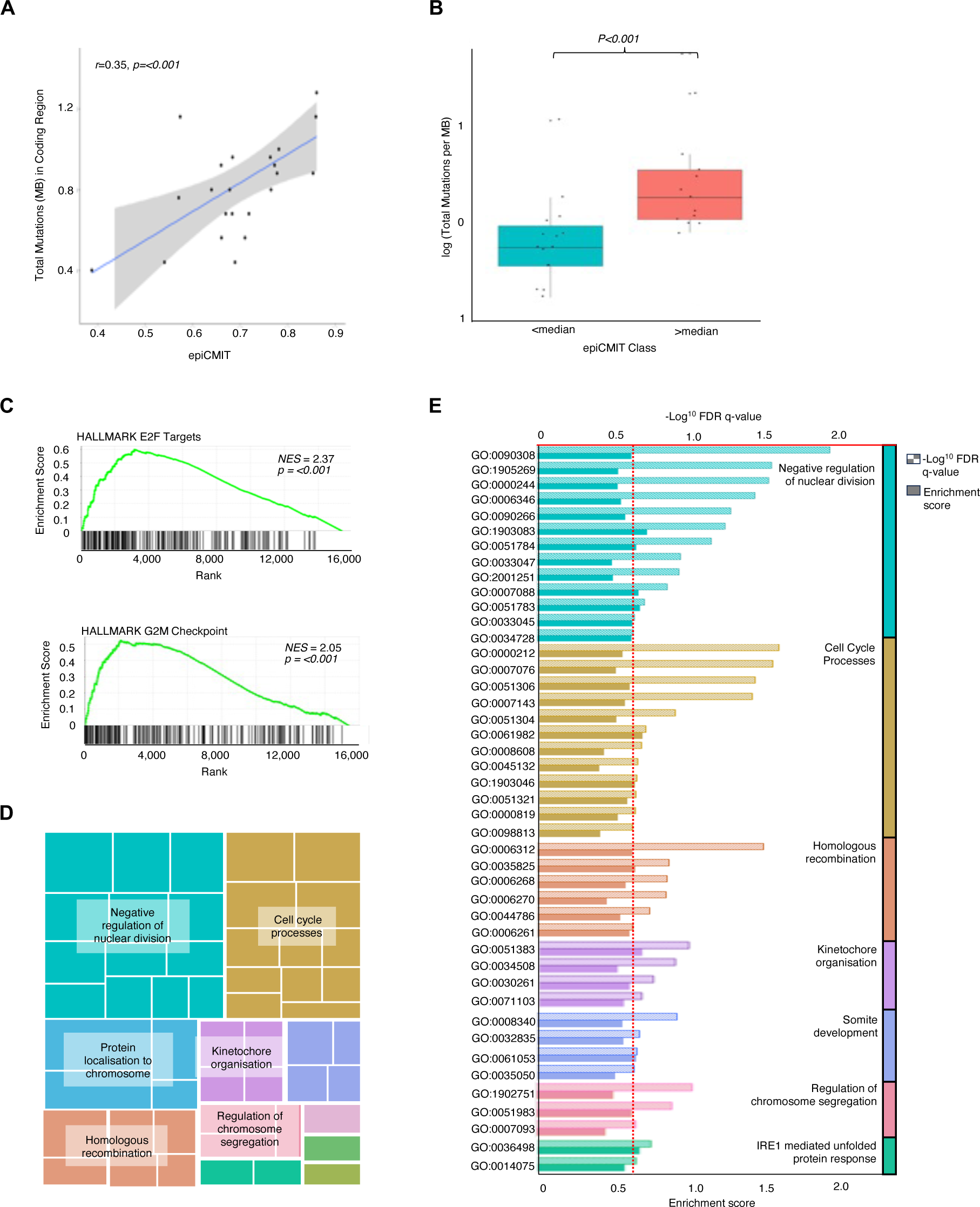
WGS and transcriptomic analysis of SMZL. **A** Scatterplot showing the significant positive correlation between the epiCMIT score and the tumor mutational burden (TMB) per Mb in coding regions. The linear regression model is depicted by the black line, and the grey area indicates the confidence intervals. **B** Box and whisker plot showing the significant difference in TMB per Mb, in patients with epiCMIT scores below and above the median (0.74) score. **C** GSEA enrichment plots for E2F target and G2M checkpoint genes, showing the position of genes assigned to these HALLMARK pathways in the ranked gene list (black bars). The enrichment scores for each gene are plotted as a density plot (green line). **D** Revigo TreeMap where each rectangle represents a GO biological process. These processes are then clustered into related terms, or branches, identified by their different colors. The size of each rectangle reflects the *p*-value. **E** Bar chart showing the significantly over-represented Gene Ontology biological process terms in the ranked gene list (FDR q value <0.25, p=<0.05). Terms are assigned to the highest branch as per the Revigo TreeMap and indicated by the colored bars to the right and ranked by the FDR q-value (cutoff of <0.25 indicated by dotted red line). Hatched bars indicate the FDR q-Value and solid bars show the enrichment score.

Next, we compared continuous epiCMIT values to other key clinico-biological features of SMZL, available for our cohort. TL data was available on 114 cases and ranged from 2.83-7.57 kb (median 3.1 kb). Interestingly we observed a significant negative correlation between epiCMIT and TL (r=-0.29, *p=*0.001) supporting the association between cell proliferation and concomitant telomere attrition (**Fig2Ci, ii**). Importantly patients with higher epiCMIT scores were significantly more likely to be female (*p*=0.02) and harbor the IGHV1-2*04 gene (*p*<0.001) with intermediate levels of IGHV SHM (97-99.9%, *p*=0.04). We also show enrichment of previously defined high-risk subgroups (AH (*p*<0.001) and NNK (*p*=0.017)) in cases with high epiCMIT (**Fig 2D**) (15, 16). Furthermore, recurrent CNAs gain(12q) (*p*=0.001) and del(7q) (*p*=0.019) (**Fig2E**), and driver somatic gene mutations, including those in *KLF2* (*p*<0.001), *NOTCH2* (*p*=0<0.01), *TP53* (*p*=0.01) and *KMT2D* (*p*<0.001) (**Fig 2F**) were associated with high epiCMIT scores. Notably, patients with low epiCMIT scores were enriched for trisomy 12 (*p*=0.003) (**Fig2E**) and *MYD88* variants (*p*=0.025) (**Fig 2F**). We did not observe a significant difference in epiCMIT in cases scored as either A or B using the HPLL scoring system for SMZL (31).

Using whole-genome sequencing data (n=23), we found that the TMB demonstrated a significant positive correlation with epiCMIT (r=0.35, p<0.001) when both were assessed as continuous variables (**Fig3A**). This was confirmed in a subgroup analysis between two patient subgroups with epiCMIT scores above (TMB=0.78) and below (TMB=0.66) the median (0.74) (*p=*<0.001) (**Fig3B**). A focused analysis of levels of SHM burden, defined by mutations within the IG loci, again showed a positive correlation with epiCMIT score (r=0.23, *p*=0.01).

In cases with mRNA-seq data (n=15), we next investigated the relationship between epiCMIT and the expression of specific genes or pathways that might be associated with a proliferative or pro-survival cellular phenotype. A mean of 2.58×10^7^ reads (range 1.5×10^7^-4.9×10^7^) per sample were generated, with 93.02% mapping to hg38 and 26865 total HGNC annotated genes detected. 15,841 genes passed filtering. Employing GSEA, using KEGG, BIOCARTA, IMMUNE and HALLMARK gene sets from MSigDB, we demonstrate enrichment of two HALLMARK pathways associated with elevated cell division: E2F target genes (NES=2.37, *p*<0.001) and the G2M checkpoint (NES=2.05, *p*<0.001) (**Fig3C**). Looking at GO terms overrepresented in differential gene expression data we extend this observation showing enrichment of terms including chromosome segregation, cell cycle processes and DNA replication (**Fig3D-E**). Finally, we employed a likelihood ratio test approach, with false-discovery correction, to identify 36 and 12 differentially over- and under-expressed genes, respectively, where the continuous range of gene expression and epiCMIT showed a significant correlation (FDR <0.05, log_2_ FC>3) (**Table S5**).

### EpiCMIT is associated with outcome in univariate and multivariate models

To investigate the prognostic significance of clinical and molecular features on TFS and OS, univariate (UV) Cox regression analysis was conducted. Among the 48 features analyzed, epiCMIT score as a continuous variable, was a highly significant predictor of shorter TFS (HR=30.6, *p*=0.001). Additional features associated with shorter TFS included gain(3q), HPLL B score, *TP53* mutation, IGHV1-2*04 usage, AH, short telomeres and female sex. (**Fig 4A**). When epiCMIT scores were multiplicatively rescaled (x10 scale factor), incremental increases of 0.1 in epiCMIT were associated with a 40% increase in risk (HR=1.4, *p*=0.001). In Kaplan-Meier analysis, using a maxstat rank statistics-based cutoff for high and low epiCMIT score, we showed that higher epiCMIT (>0.76) was associated with significantly shorter TFS (median 3 vs. 23 months) (*p*=<0.001) (**Fig 4B**). Additionally, a significant increase in the proportion of patients requiring therapeutic intervention was observed in association with higher epiCMIT scores (*p*=0.003) (**Fig 4C**).

**Figure 4.**
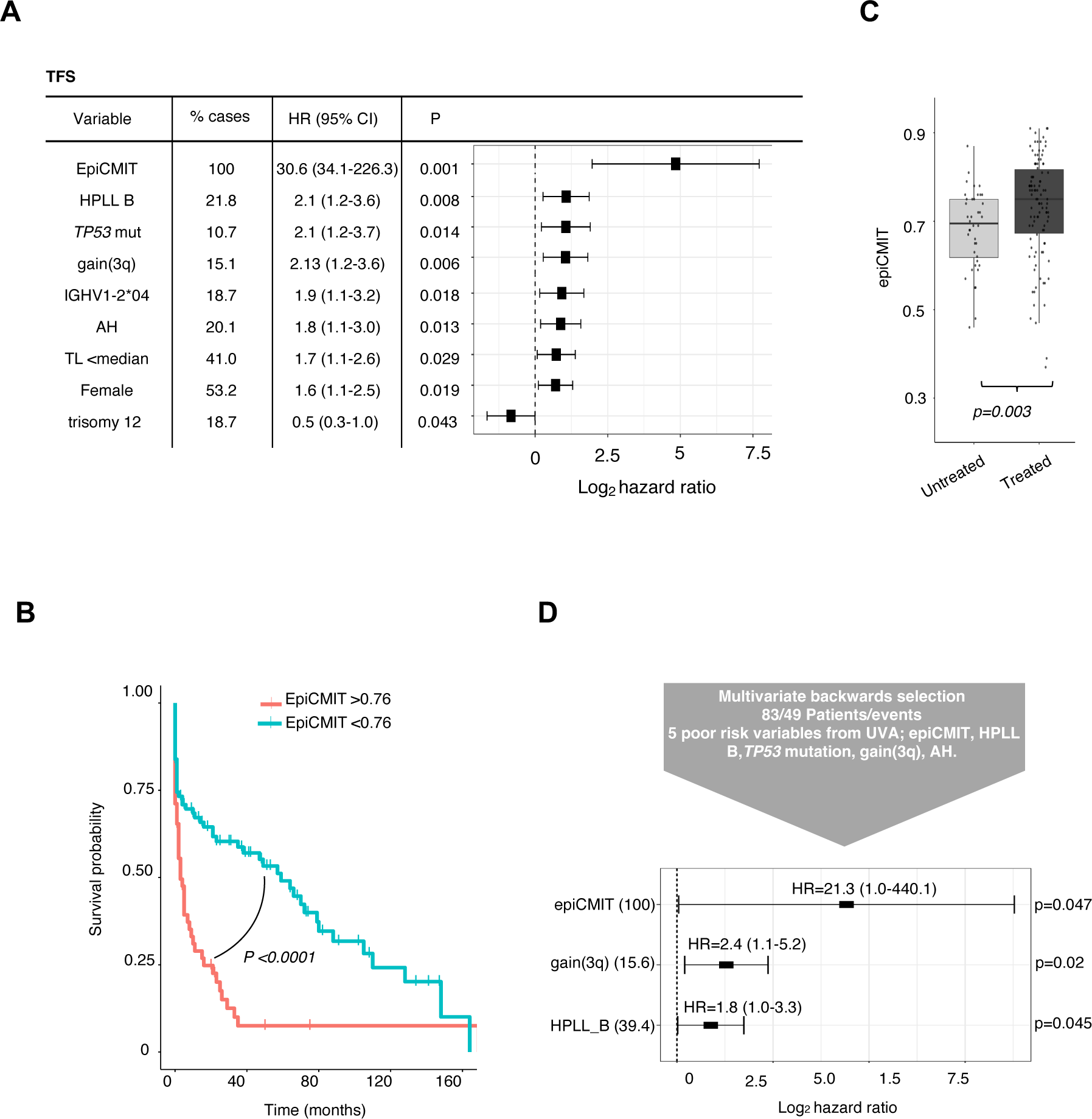
Clinical outcome in SMZL is associated with epiCMIT. **A** Forest plots showing the hazard ratios and confidence intervals for variables significant (p<0.05) for TFS, in univariate (UVA) cox regression analysis. The proportion of cases going into the analysis that are positive for each variable is shown. **B** Kaplan Meier curves for TFS (months) with high or low epiCMIT assigned according to the maxstat rank statistics-based cutoff (0.76) (pink line and blue lines, respectively). **C** Box and whisker plot showing the significant difference in epiCMIT scores in treated compared to untreated patients. Mann-Whitney U test with Benjamini-Hochberg correction was used to determine statistical significance (p=0.05). **D** Forest plots showing the variables significant for TFS in multivariate (MVA) Cox Proportional Hazards models (backwards selection). The proportion of cases going into the analysis that are positive for each variable is shown ().

In our diverse treatment cohort, where lymphoma-unrelated deaths are common, epiCMIT as a continuous variable was not an indicator of shorter OS in UV analysis (**Fig S4A**). However, Kaplan-Meier analysis showed that higher epiCMIT (>0.72, maxstat rank statistics-based cutoff) was significantly associated with shorter OS (median 63 vs. 68 months) (*p*=0.031) (**Fig S4B**), and increased mortality was observed in patients with high epiCMIT (*p*=0.004) (**Fig S4C**).

To establish a prognostic model, a multivariate Cox Proportional Hazards model, using backwards selection, was constructed for TFS. The model incorporated the five most significant variables, as determined by UV analysis, ranked according to p-value. It was built using data from 83 patients with complete data, encompassing a total of 49 events. EpiCMIT as a continuous variable maintained a highly significant independent prognostic impact in the context of the HPLL clinical scoring system, *TP53* mutation, gain(3q) and the methylation-based classifier, AH. Overall, epiCMIT, gain(3q) and HPLL B score are the most significant independent variables associated with TFS in SMZL (**Fig 4D**).

## Discussion

In this study, we undertake an investigation into the clinico-biological significance of epiCMIT in SMZL. Our transcriptomic findings lend support to the concept that SMZL tumor cells exhibiting elevated epiCMIT levels possess the potential for subsequent cellular division, a phenomenon demonstrated in other B-cell neoplasms (17). In SMZL, this cellular activity is facilitated through the activation of mitotic, DNA replication, and metabolic pathways. Employing comprehensive clinical, biological, and (immuno)genomic features within robust survival models, we illustrate the efficacy of epiCMIT as a prognostic indicator in SMZL. This identification proves valuable in discerning high-risk patients and predicting diminished treatment-free survival, thereby emphasizing the potential utility of epiCMIT in guiding clinical decision-making. In MVA, both epiCMIT and chromosome 3q gain emerge as independent markers of TFS. The latter event leads to the duplication of the proto-oncogene *BCL6*. The occurrence of gain(3q) has previously been associated with the transformation of SMZL to DLBCL (32), and was observed in 22.2% of our cases that eventually underwent transformation. We noted no substantial differences in epiCMIT scores between samples derived from splenic and peripheral blood cells, indicating the informativeness of epiCMIT in both sample types. Furthermore, peripheral SMZL cells retain the methylation fingerprint associated with cell division in splenic proliferative niches.

We have illustrated that epiCMIT exhibits high levels in patients carrying well-established recurrent CNAs such as del(7q) and gain(12q), along with gene mutations (e.g., *NOTCH2, TP53*, and *KLF2*). The clinical impact of these genomic lesions is likely attributed to increased proliferation in these cases, especially given the independent significance of epiCMIT in MVA models of TFS that incorporate genomic information. These findings suggest that, like other tumors, specific genomic defects in SMZL may undergo positive selection, conferring a heightened proliferative advantage. Furthermore, we identified a second subgroup of patients characterized by a low epiCMIT score, trisomy 12, and *MYD88* mutations. While epiCMIT exhibited a significant association with TFS, it did not emerge as an independent marker of OS. It is important to note that OS is often confounded by the diversity of treatment regimens in SMZL and deaths from unrelated causes in an elderly patient cohort.

Through telomere length (TL) analysis, we demonstrated a modest yet significant negative correlation with epiCMIT, supporting the clinical importance of this biomarker and the idea that cellular proliferation persists alongside telomere attrition. While various studies have explored the clinical relevance of TL in other mature B-cell tumors (22, 33–35), our study represents the first investigation of TL in SMZL, including a comparative analysis of the clinical value of each biomarker. Our analysis suggests that epiCMIT, accumulated gradually and consistently during B cell differentiation, emerges as a more clinically relevant biomarker compared to TL. Unlike TL, which does not exhibit a linear decrease and instead shows a marked increase in germinal center-derived B-cells due to transient telomerase upregulation (36), epiCMIT’s consistent accumulation makes it a more robust biomarker. Like chronic lymphocytic leukemia (CLL), proliferation leading to telomere shortening may provide SMZL cells with a survival advantage by inducing genomic instability, driving further clonal evolution and disease progression (37, 38). The association we observed between elevated epiCMIT and both female sex and IGHV1-2*04 usage warrants future investigation. This may be linked to the higher prevalence of autoimmunity in females (39) and in SMZL cases in general (12, 40), and the fact that IGHV1-2*04 encoded B-cell receptors are most likely autoreactive and chronically activated by autoantigen (41).

Arribas and colleagues conducted the sole other DNA methylation study on SMZL, while Bonfiglio and colleagues proposed a genetically defined classification system for SMZL. Our study holds the advantage of a substantial patient cohort, a significant enhancement in the resolution of DNA methylation profiling technology, and more extensive clinico-biological, genomic, and transcriptomic data. Both previous studies identify a group of high-risk patients—High-M and NNK—associated with recurrent genomic features such as del(7q), IGHV1-2*04, and *NOTCH2* mutations. While High epiCMIT scores identified both the High-M and NNK subgroups, our study demonstrated superior performance in predicting shorter TFS in our cohort. This suggests that epiCMIT captures additional biological factors associated with an unfavorable patient outcome. Duran-Ferrer *et al* propose that the clinical utility of epiCMIT should be considered only in the context of tumors with the same ground-state proliferative history as the normal cell of origin. They most convincingly demonstrated this in CLL, where epiCMIT provided valuable clinical information when accounting for the three CLL epitypes with distinct cell origins. In contrast, our study reveals the broad clinical relevance of epiCMIT across all SMZL cases, potentially indicating a more similar ground-state level of non-malignant proliferation in SMZL compared to other B-cell tumors.

Future studies employing high-resolution epiCMIT analysis, potentially at the single-cell level, could explore the dynamic evolution and sub-clonal architecture of SMZL, tracing cells with proliferative potential back into early-stage disease and providing innovative methods for predicting future disease onset. Moreover, investigating the utility of epiCMIT in specific therapies, particularly rituximab-based regimens administered in clinical trials, may aid in predicting patients destined to respond favorably to certain treatments. Our analysis contributes to a growing body of evidence suggesting that methylation profiling serves as a valuable clinical tool in the diagnostic workup of lymphoma patients. For instance, future platforms could offer methylation-based tumor classification, epiCMIT quantification, exon-level copy number detection, and mutational analysis in a single, cost-effective assay system.

The key strengths of our study lie in the exhaustive genome-wide methylation analysis of a substantial cohort affected by a rare lymphoma, complemented by expansive clinical, biological, and (immuno)genetic data. To ensure diagnostic accuracy, particularly for historical cases diagnosed before the currently accepted criteria, we limited our analysis to cases from centers specializing in SMZL. While the rarity of SMZL prevented the inclusion of a validation cohort, the support for the significance of epiCMIT in SMZL comes from previously published analyses of other B-cell tumors (17). Despite demonstrating a robust impact of epiCMIT on TFS, our assessment of overall survival (OS) faced limitations due to varied treatment modalities and the prevalence of elderly SMZL patients succumbing to unrelated causes. Furthermore, the limited data in our cohort prevented an evaluation of epiCMIT’s effectiveness in predicting transformation to large cell lymphoma.

In summary, our extensive analysis of a large SMZL patient cohort has unveiled fresh insights into the biological implications of methylation in SMZL. We identified a high-risk subgroup, characterized by a shorter treatment free survival (TFS), distinguishable by a high epiCMIT score-as an independent prognostic biomarker. This subgroup aligns with established high-risk features, including del(7q), IGHV1-2*04, and mutations in key driver genes. These findings enrich our comprehension of SMZL pathogenesis and hold potential for guiding more precise clinical management strategies for affected individuals.

## Data Availability

All data produced in the present study are available upon reasonable request to the authors

## Acknowledgments

This work was supported by research grants from the Kay Kendall Leukemia Fund (873, 1104), and Cancer Research UK (ECRIN-M3 accelerator award C42023/A29370). We acknowledge the use of the IRIDIS High Performance Computing Facility, and associated support services at the University of Southampton. We thank the University of Southampton Faculty of Medicine Tissue Bank for support in the completion of this work. MDF is supported by a postdoctoral grant from the Spanish Association Against Cancer. R.R is supported by grants from the Swedish Cancer Society, the Swedish Research Council, Region Stockholm, and Radiumhemmets Forskningsfonder, Stockholm. Sequencing was performed by the SNP&SEQ Technology Platform in Uppsala. The facility is part of the National Genomics Infrastructure (NGI) Sweden and Science for Life Laboratory. We thank the Genomics and Proteomics Core Facility of the DKFZ (Heidelberg, Germany) for excellent support.

## Authorship Contributions

HP, DB, MRZ, DO, SE & JS designed the study. HP, CJO, MRZ, HA, LC, SS, MS, MP, ZD, DW, CP, NMB & DW performed laboratory work. HP, DB, AM, LB, JT, CJO, VL & BS performed data analysis. HP, DB, DO & JS wrote the manuscript. FF, RW, KS, AX, EK, RR, LS, PG, CT, CK, GP, DR, SW, MA, AF & MS, RMA provided study material and clinical data. JG, FS, LK, CO, RR, RW, MDF, JIMS provided expert advice and all authors reviewed the manuscript.

