## Supplementary Methods for "Proliferative History Is a Novel Driver of Clinical Outcome in Splenic Marginal Zone Lymphoma"

### Patient Cohort

Our cohort comprises 142 primary SMZL patients, all meeting established diagnostic criteria (splenic histology, immunophenotype, lymphocyte morphology, IGHV and cytogenetic / FISH data). In non-splenectomized (n=85/142) cases that met the baseline criteria for SMZL diagnosis, supporting data was considered including IGVH1-2*04 usage (derived from standard diagnostic tests), del(7q) (derived from DNA methylation analysis), *NOTCH2* and *KLF2* mutations. Informed consent was obtained from all patients in accordance with the Helsinki declaration, and the study was approved by our regional and local ethics committees. Prior to DNA extraction (DNeasy blood and tissue kit, Qiagen, Hilden, Germany), the CD19+/CD45- SMZL cells were purified from the peripheral blood or spleen cells using the EasySep Human B Cell enrichment kit without CD43 depletion (StemCell Technologies, Cambridge, UK). Tumor purity of greater than 85% was confirmed in all cases by flow cytometry (CD19).

### DNA Methylation analysis

142 DNA samples were processed, as previously reported (1, 2) using the Illumina Infinium Human Methylation 450 BeadChip (n=111) and Illumina Infinium Methylation EPIC BeadChip (n=31) (Illumina, Hayward, CA, USA), according to manufacturer’s instructions, at the Genomics and Proteomics Core Facility of the DKFZ (Heidelberg, Germany). DNA processing was performed using RnBeads v2.93 (RRID:SCR_010958) from raw intensity data through to import, annotation against hg19, quality control, SWAN normalization, differential methylation analysis and output beta/M-values. As we had a mix of 450k and EPIC array data, an intersected CpG list was generated for CpGs shared by both platforms (n=428,568). Following removal of SNP enriched probes and unreliable probes using the GreedyCut method (n=9,778), the beta values from 418,790 CpGs were used for downstream analysis. Tumor purity estimates, calculated from DNA methylation data (3), confirmed FACS based purity scores in all samples. Downstream analysis was three-fold; generating copy number data, epiCMIT scores and the recapitulation of the high- and low-global methylation groups previously defined by Arribas and colleagues (4). Conumee (5) was used to produce profiles of copy number alterations (CNA) from mean intensity signals, which were manually curated; with the exception of a single 0.85Mb deletion of *ATM* and three 0.8Mb deletions of the Chr 13q14 *DLEU2* locus, sub-chromosomal gains or losses were only included if they were ≥1 Mb, were defined by a minimum of 5 consecutive probes and had segmented means of +/-0.33 (log_2_ ratio of probe intensity). CN calls were compared with WGS-based calls where data was available, with 100% concordance. Genomic complexity was defined as the presence of ≥3 CNA. EpiMIT-hypo and -hyper scores were calculated as per Duran-Ferrer *et al* (6) using the Estimate.epiCMIT function published by the authors. For each patient, the highest score from the epiCMIT-hypo and -hyper scores was selected to derive a unique epiCMIT value (termed epiCMIT). Our intersected 450k/EPIC methylation data covered 174/184 and 1069/1164 of the hypermethylated and hypomethylated CPGs in this signature, respectively. Cases were assigned to the High (n=28) or Low (n=114) epigenetic subgroup as per Arribas *et al* by clustering beta values for the CpGs described as the top100 most variable, of which 86/100 were represented in our intersected 450k/EPIC array data (4) (**Table S1**). Clustering was performed using Euclidean distance and complete linkage.

### Mutation detection with targeted re-sequencing

DNA samples from 133 SMZL cases were analyzed with a bespoke Agilent Haloplex Target Enrichment system as previously described (7), using a panel designed with SureDesign (<https://earray.chem.agilent.com/suredesign/>) to enrich 383.74 kb of genomic DNA for 62 genes and genomic regions, selected for their clinical relevance to SMZL and other B-cell malignancies (**Table S3**), resulting in 98.95% *in silico* coverage of selected regions. Matched germline material was not available so high confidence variants were identified using a bespoke bioinformatics pipeline and filtering strategy; FASTQ files were run through FastQC for quality control, and through Surecall Trimmer v4.0.1 to identify and remove adaptor sequences and trim low quality reads (qual < 20). BWA-MEMv0.7.12 was used to align reads to the hg38 reference genome using default options. Samtools v1.2.3 converted SAM files into BAMs and Picard v2.8.3 sorted and indexed the BAMs. Duplicate reads were marked and merged using LocatIt v4.0.1 learning all possible start/stop combinations as it was reading the data, rather than using the BED file. Resulting BAMs were sorted and converted into FASTQ by Samtools and Picard respectively. Pear v1.97 merged the paired end reads (FASTQs). The output from Pear were assembled, and unassembled FASTQs were then run through BWA-MEM and aligned to the hg38 reference genome. BAMs were again converted to SAM format (Samtools) and sorted by Picard. Picard merged the assembled and unassembled BAMs and the resulting merged BAM was run through GATKs Haplotype caller (v3.7) which called variants via local re-assembly of haplotypes. The resulting VCF files were annotated using Annovar software (v2016Feb01) with the following databases: The Genome Aggregation Database77, 1000Genomes Project78, NHLBI GO Exome Sequencing Project79, Exome Aggregation Consortium77, Kaviar, Haplotype consortium, dbsnf33a, ClinVar, COSMIC, nci60 and an in-house SMZL reference database (SMZLrefDB).

After annotation, variants were filtered to enrich for high confidence somatic mutations. Initial filtering excluded intronic and intergenic variants (except for *NOTCH1* and *NOTCH2* 3’-UTRs and *PAX5* non-coding variants), variants with a frequency > 1% in databases of known germline variation, and variants with a total depth less than 30. This allowed 95% confidence in identifying variants with VAFs as low as 0.10. To determine the likelihood of a variant being pathogenic and exclude germline variation, variants were annotated using CADD phred scores (8), SpliceAI scores (9) and the mutation significance cut-off (MSC)(10) for both the Human gene Mutation Database (HGMD) (11) and ClinVar (12) with a 90% confidence interval. Variants with a CADD Phred score ≥ 15 or ≥ MSC cut-offs were excluded as well as those with a SpliceAI score < 0.5. Lastly synonymous mutations were excluded. Variants passing filter were manually curated in Integrative Genomics Viewer (IGV) according to the standard operating procedure described by Barnell *et al* (13).

As described by Bonfiglio *et al* (14), the presence of somatic mutations in a set of 14 genes, can be used to assign SMZL patients into 2 prominent clusters termed NNK (NF-Kb, NOTCH and KLF2 modules) and DMT (DNA damage response, MAPK and TLR modules). Except for *PTPN11*, all genes used by the authors in this classification model were included on our Haloplex panel and were used to classify patients as NNK (n=52) or DMT (n=35). Patients lacking mutations in the 13 key genes were assigned to an ‘unclassified’ group (n=55) (**Table S1**).

### Telomere length analysis:

Telomere length (TL) relative to a standard reference sample (K562 cell line) was determined using monochrome multiplex PCR (MMQ-PCR) as previously described (15), in 114 patients. Absolute TL in kb was extrapolated, using linear regression, from 82 CLL cases with STELA data; previously published data (15) shows a high concordance (Spearman correlation 0.80) between 111 CLL4 patients analyzed by both MMQ-PCR and STELA. Using the K562 standard, MMQ-PCR was repeated for 82 of these cases, and a correlation of 0.816 was achieved. Patients were classified as having long or short telomeres based on the median length (3.12 kb).

### Whole-genome, transcriptome and microRNA sequencing

Twenty-three patients, with diverse epiCMIT scores and viable cells available, were processed with WGS (n=23/23), mRNA-Seq (n=15/23). For WGS DNA was extracted from tumor B-cells (DNeasy Blood and Tissue Kit, Qiagen) and matched saliva (Oragene DNA Saliva kit, DNA Genotek, Ottawa, Canada), prior to library preparation with the Illumina TruSeq Nano Kit, and paired-end sequencing (2x150bp) with 30x coverage on the Illumina HiSeqXTen system at the SNP&SEQ Technology Platform, Science for Life Laboratory at Uppsala University, Sweden. QC of FASTQ files was performed using FastQC v0.11.9 and MultiQC v1.11. The GATK Best Practices Workflow (16) or ‘Data Pre-Processing for Variant Discovery’ was utilized for read alignment and processing. BWA-MEM v0.7.17 was used to align reads to the hg38 reference genome. Picard v2.18.14 was used to convert SAM files into BAM files, mark duplicate reads and resort and index the BAMs. Base score recalibration was then performed using GATK software. QC of the aligned BAMS were performed using QualiMap v2.2.1.

The GATK Best Practices ‘Somatic short variant discovery (SNVs + Indels)’ pipeline was applied to our dataset. The MuTect2 module v4.2.2 was used to call somatic SNVs in individual tumor-normal matched samples. The somatic variant calling pipeline first creates a ‘panel of normals’ (PoN) using all normal BAMS and containing all collated sites that were present in two or more samples, allowing for the capture of common artefactual or germline variant sites. Somatic variant candidate calling with Mutect2 implements both this PoN and a germline variant dataset derived from gnomAD v2. These analysis-ready variants were then annotated by the Functotator software v4.2.2 with the following databases: UniProt v2014_12 (The UniProt Consortium, 2017), ORegAnno v20160119 (17), gnomAD (Genome and Exome v2.1), GENCODE v34 (18), dbSNP build 151 (19), COSMIC v84 (20), ClinVar v20180429_hg38 (21), Cancer Gene Census (CGC) v.full_2012_03-15 (22) and Hugo Gene Nomenclature (HGNC) v.Nov-30-2017.

Visualization of SNV calling data from resulting .maf files was done in RStudio v1.4.1717, using the MAFTools v2.10.0 (23) and ggplot2 v3.3.5 packages. The coding tumor mutational burden (TMB) was assessed using MAFTools v2.10.0 and was defined as the burden of mutations per mega-base pair, within the coding regions of the genome.

mRNA sequencing was performed on RNA extracted using the Qiagen miRNeasy Tissue/cells Advanced micro kit (Qiagen, Hilden, Germany). mRNA libraries were prepared using the NEBNext Poly(A) mRNA Magnetic Isolation Module (NEB, Massachusetts, USA) and sequencing was performed on the Illumina NovaSeq6000. mRNA -Seq analysis was performed as previously reported (1). mRNA fasta files were aligned to the hg38 Reference genome using STAR aligner v2.7.10b and read counts were calculated through HTseq-count against Gencode GRCh38.p14 v44 GTF. Differential gene expression analysis was conducted using EdgeR v3.42.4 (RRID:SCR_012802) against epiCMIT as a continuous variable using likelihood ratio tests (significance; FDR p<0.05 with Benjamini-Hochburg correction). GSEA was performed against the Hallmark, KEGG and immune gene sets obtained from MSigDBv2023.1Hs (24-27). All analytical processes were conducted using R v4.3.1 (RRID:SCR_001905).

### Statistics

Relationships between variables, that were observed in at least 5% of cases (n=7), were compared using the Fisher’s exact and Mann-Whitney U tests (significance; *p*=0.05). Kaplan-Meier curves were used to investigate overall survival (OS) (measured from date of diagnosis to date of death from any cause or last follow-up) and treatment free survival (TFS) (measured from date of diagnosis to date of first treatment or date of death from any cause or last follow-up). Differences in survival measures between subgroups were tested with the log-rank test. Multivariate Cox Proportional Hazard models were generated for TFS using stepwise backwards elimination incorportaing data from 83 patients with 49 events. The five most significant variables, as determined by UV analysis, ranked according to p-value, were incorporated into the model. The variable with the largest p-value was eliminated until all remaining variables had a significant p-value (p<0.05). All analyses were performed in R v3.6.1 (RRID:SCR_001905) using custom code.
