## Supplementary Figures for "Proliferative History Is a Novel Driver of Clinical Outcome in Splenic Marginal Zone Lymphoma"

### Slide 1
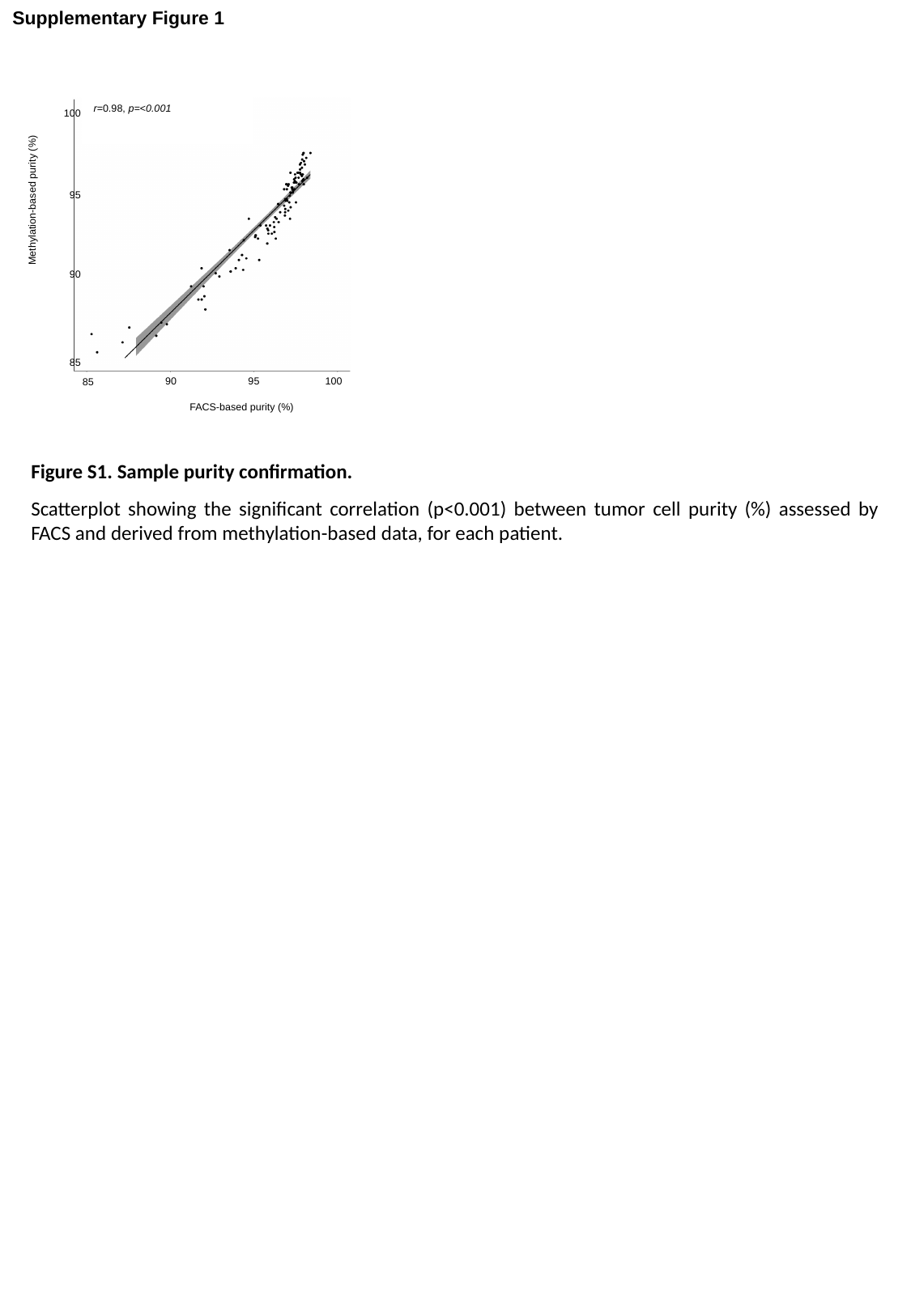

Supplementary Figure 1
r=0.98, p=<0.001
100
Methylation-based purity (%)
95
90
85
90
95
100
85
FACS-based purity (%)
Figure S1. Sample purity confirmation.
Scatterplot showing the significant correlation (p<0.001) between tumor cell purity (%) assessed by FACS and derived from methylation-based data, for each patient.

### Slide 2
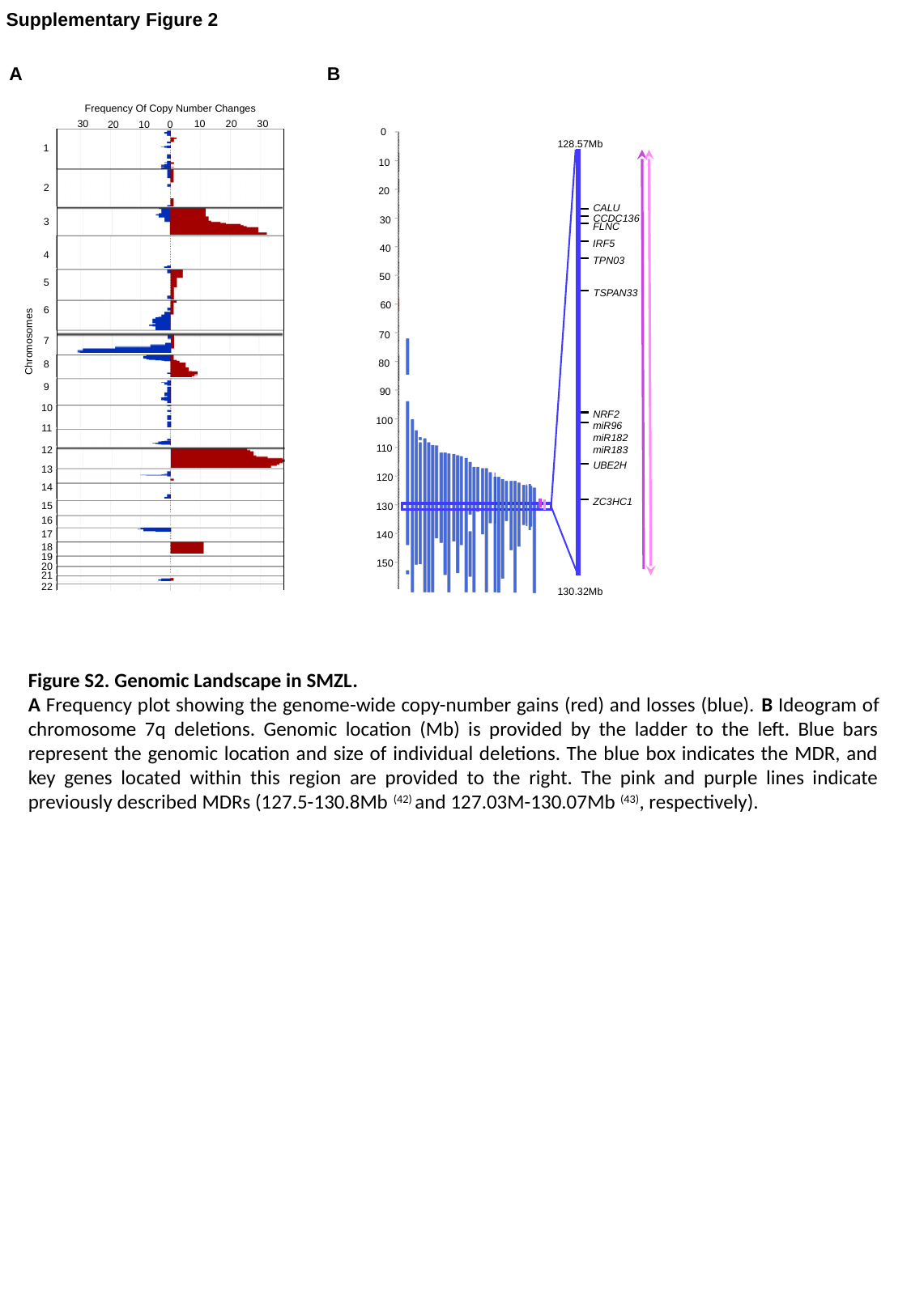

Supplementary Figure 2
A
B
Frequency Of Copy Number Changes
30
10
20
30
0
10
20
0
128.57Mb
10
20
CALU
CCDC136
30
FLNC
IRF5
40
TPN03
50
TSPAN33
60
70
80
90
NRF2
100
miR96
miR182
miR183
110
UBE2H
120
ZC3HC1
130
140
150
130.32Mb
1
2
3
4
5
6
7
Chromosomes
8
9
10
11
12
13
14
15
16
17
18
19
20
21
22
Figure S2. Genomic Landscape in SMZL.
A Frequency plot showing the genome-wide copy-number gains (red) and losses (blue). B Ideogram of chromosome 7q deletions. Genomic location (Mb) is provided by the ladder to the left. Blue bars represent the genomic location and size of individual deletions. The blue box indicates the MDR, and key genes located within this region are provided to the right. The pink and purple lines indicate previously described MDRs (127.5-130.8Mb ^(42)^ and 127.03M-130.07Mb ^(43)^, respectively).

### Slide 3
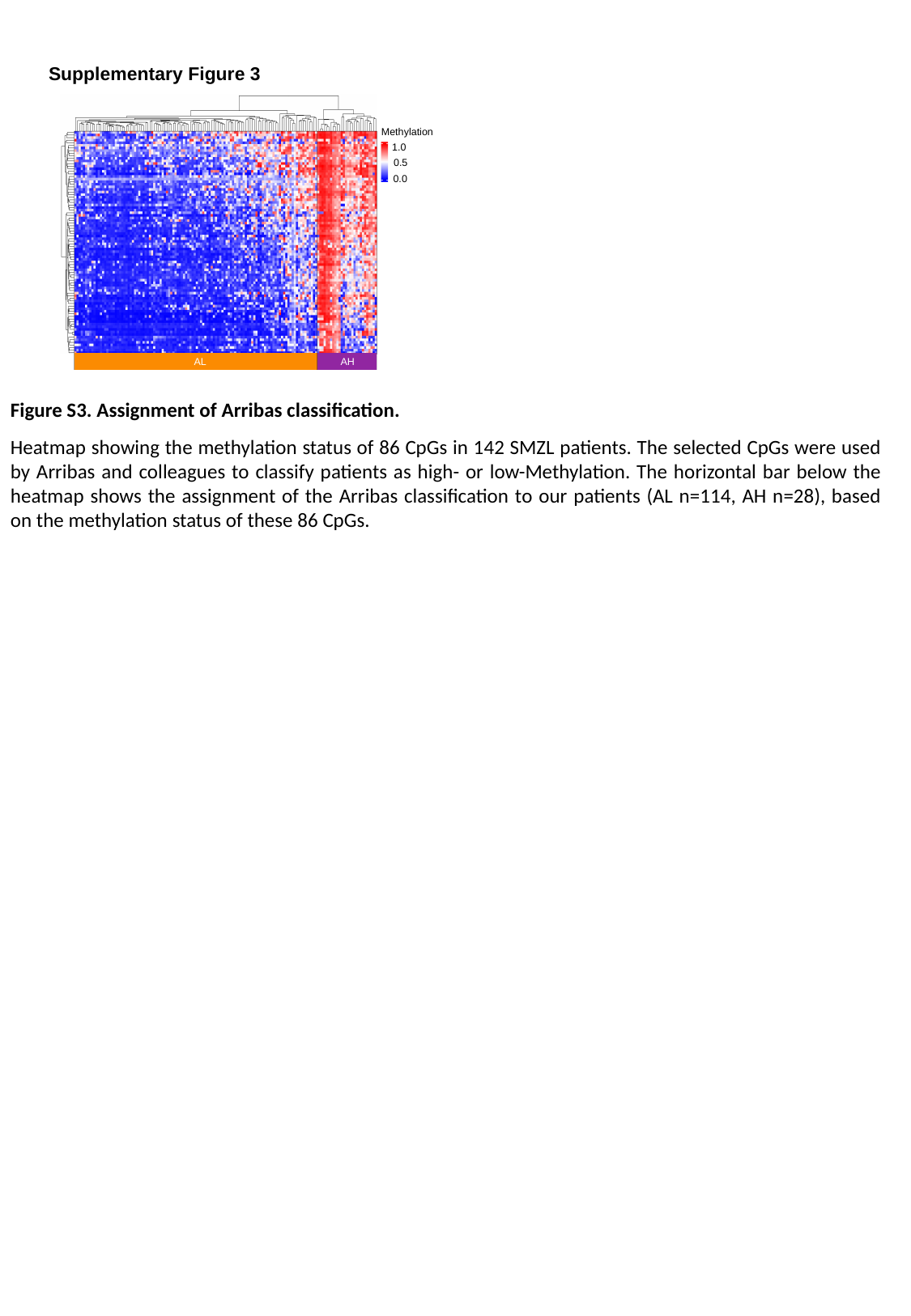

Supplementary Figure 3
Methylation
1.0
0.5
0.0
AL
AH
Figure S3. Assignment of Arribas classification.
Heatmap showing the methylation status of 86 CpGs in 142 SMZL patients. The selected CpGs were used by Arribas and colleagues to classify patients as high- or low-Methylation. The horizontal bar below the heatmap shows the assignment of the Arribas classification to our patients (AL n=114, AH n=28), based on the methylation status of these 86 CpGs.

### Slide 4
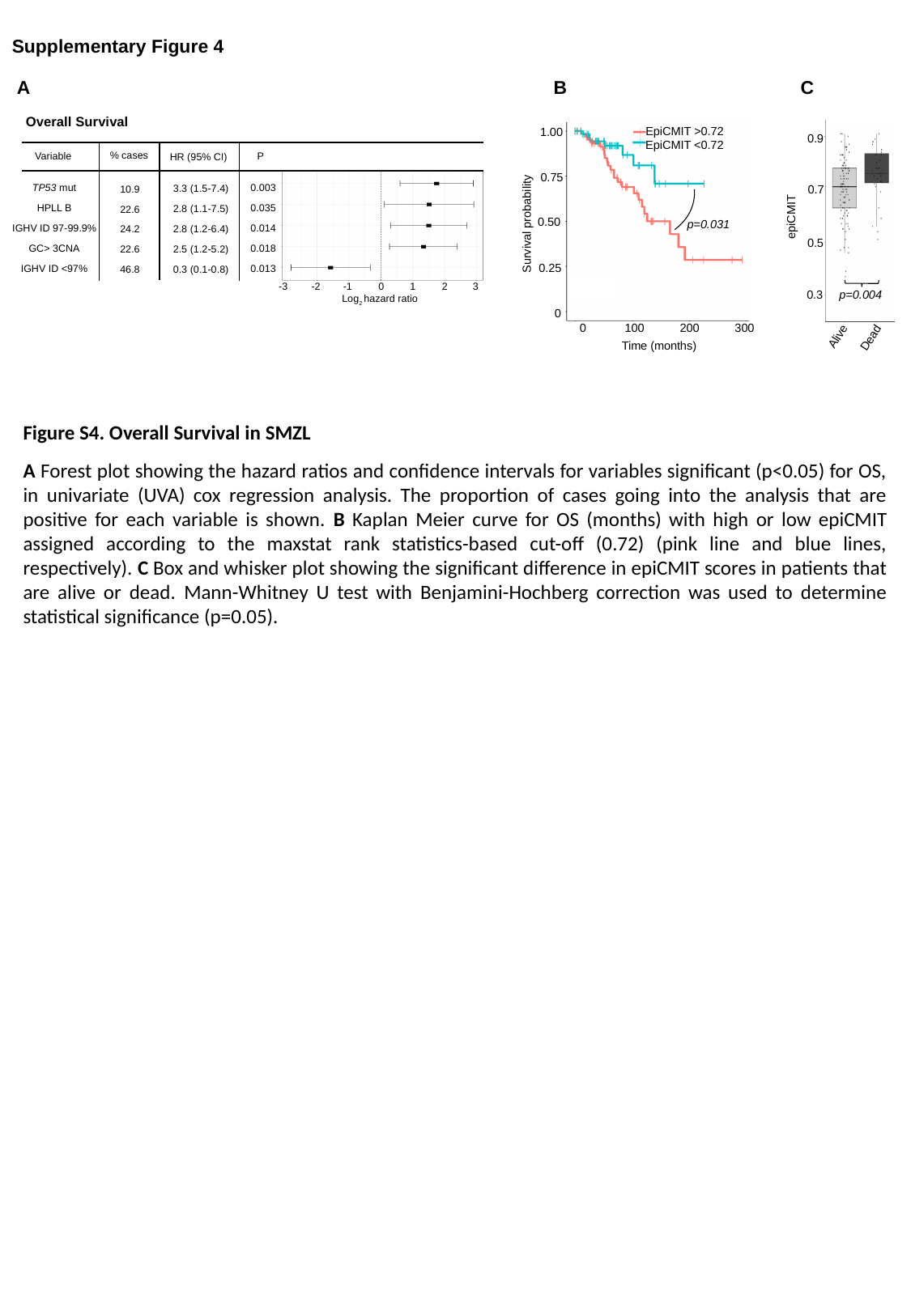

Supplementary Figure 4
B
A
C
Overall Survival
EpiCMIT >0.72
1.00
EpiCMIT <0.72
0.75
0.50
p=0.031
Survival probability
0.25
0
0 100 200 300
Time (months)
0.9
0.7
epiCMIT
0.5
0.3
p=0.004
% cases
Variable
P
HR (95% CI)
TP53 mut
HPLL B
IGHV ID 97-99.9%
GC> 3CNA
IGHV ID <97%
0.003
0.035
0.014
0.018
0.013
3.3 (1.5-7.4)
2.8 (1.1-7.5)
2.8 (1.2-6.4)
2.5 (1.2-5.2)
0.3 (0.1-0.8)
10.9
22.6
24.2
22.6
46.8
1
2
-1
0
-3
-2
3
Log_2_ hazard ratio
Alive
Dead
Figure S4. Overall Survival in SMZL
A Forest plot showing the hazard ratios and confidence intervals for variables significant (p<0.05) for OS, in univariate (UVA) cox regression analysis. The proportion of cases going into the analysis that are positive for each variable is shown. B Kaplan Meier curve for OS (months) with high or low epiCMIT assigned according to the maxstat rank statistics-based cut-off (0.72) (pink line and blue lines, respectively). C Box and whisker plot showing the significant difference in epiCMIT scores in patients that are alive or dead. Mann-Whitney U test with Benjamini-Hochberg correction was used to determine statistical significance (p=0.05).
